# Real-World Cardiovascular Outcomes with a Carbohydrate-Reduced Telemedicine Intervention

**DOI:** 10.1101/2025.10.27.25338916

**Authors:** Priya V. Shanmugam, Caroline G.P. Roberts, Shaminie J. Athinarayanan, Adam J. Wolfberg, Stephen Raskin, Francisco Lopez-Jimenez

**Author notes:** Address for correspondence: Caroline G.P. Roberts, MD Virta Health, 3513 Brighton Blvd, Suite 310, Denver, CO 80216. Data Sharing Statement: The data underlying this article was provided by the third party, Komodo Health, under license and cannot be shared publicly. The source data for this study were licensed by Virta Health from Komodo Health, and hence may not be shared publicly.

## Abstract

**Importance:** Carbohydrate-reduced nutrition improves multiple metabolic risk factors, but its influence on long-term cardiovascular (CV) outcomes remains uncertain.

**Objective:** To evaluate the impact of an individualized telemedicine nutrition therapy on the incidence of CV events.

**Design:** Claims-based cohort study of adults with type 2 diabetes or obesity receiving telemedicine nutrition therapy versus a propensity score matched control group between January 1, 2016 and June 1, 2025, with a median follow-up of approximately two years.

**Setting:** The intervention cohort, a US-based nationwide digital health clinic, received individualized telemedicine nutrition and clinical care. The matched control cohort was derived from a commercial claims database.

**Participants:** Adults with type 2 diabetes or obesity and no CV events one year prior to index date.

**Exposure:** Individualized nutrition therapy (INT) program combining telemedicine clinical care and carbohydrate-reduced nutrition counseling.

**Main Outcomes and Measures:** Four prespecified primary outcomes included 1) 3-point major adverse CV events (MACE), defined as nonfatal myocardial infarction, nonfatal stroke, or death from any cause; 2) 6-point MACE, adding percutaneous coronary intervention, hospitalization for heart failure or unstable angina; 3) all new-onset CV events; and 4) new-onset hypertension. Secondary outcomes included all-cause mortality, and safety outcomes related to arrhythmias.

**Results:** In 4,877 participants in each cohort (mean age, 51 [SD 9.5] years; 2,939 [60.3%] female), INT was associated with reduced risk of all primary outcomes. Incidence per 1,000 person-years was 4.1 vs 9.3 for MACE-3, 5.7 vs 10.8 for MACE-6, 27.7 vs 36.9 for all new onset CV disease, and 41.8 vs 49.3 for new-onset hypertension. Hazard ratios were 0.44 (95% CI, 0.29-0.65; P <0.001) for MACE-3, 0.52 (95% CI, 0.37-0.73; P <0.001) for MACE-6, 0.70 (95% CI, 0.59-0.82; P <0.001) for all CV disease, and 0.81 (95% CI, 0.70-0.93; P <0.001) for new-onset hypertension.

**Conclusions and Relevance:** Individualized telemedicine nutrition therapy was associated with lower CV event incidence compared to controls, suggesting the intervention may confer cardioprotection.

**KEY POINTS:** *Question:* What is the impact of an individualized telemedicine nutrition program on cardiovascular events?

*Findings:* In this claims-based cohort study, individuals receiving carbohydrate-reduced nutrition therapy had significantly reduced risk of all primary outcomes compared with matched controls: 56% lower 3-point MACE, 48% lower 6-point MACE, 30% lower risk of all new-onset cardiovascular events, and 19% lower new-onset hypertension. The risk of all-cause mortality was also directionally reduced.

*Meaning:* Telemedicine nutrition therapy was associated with favorable cardiovascular outcomes compared to usual care.

## INTRODUCTION

Cardiovascular disease is the leading cause of death worldwide (1), and accounts for one-third of all mortality in the United States (2). Although Mediterranean diet interventions and several drug classes effectively reduce CV events, substantial residual risk remains (3), particularly among individuals with type 2 diabetes (T2D) and obesity. Given the rising prevalence of these conditions (4), there is an urgent need to identify strategies that address underlying metabolic dysfunction, improve glycemic control, and mitigate long-term CV risk.

We examined outcomes of a digitally delivered, continuous remote care model integrating individualized nutrition therapy (INT) focused on carbohydrate reduction to sustain nutritional ketosis, diabetes medication management, and ongoing health coaching, delivered through a continuous remote health platform. Clinical trials and real-world studies of the INT program demonstrate broad cardio-kidney-metabolic health improvements including weight loss, improved glycemic control, decreased diabetes medications, and durable reductions in systemic inflammation and visceral fat (5–11). Comparable findings have been reported by other investigators using carbohydrate-reduced and ketogenic nutrition approaches (12, 13). Yet, concerns persist that these improvements may be counterbalanced by rises in LDL-cholesterol (LDL-C), prompting debate about the net impact on CV risk (14).

Given these demonstrated cardiometabolic benefits and the ongoing discourse surrounding LDL-C, there is a pressing need to determine how this approach affects CV event rates, especially in real-world clinical settings. Using longitudinal medical and prescription claims data we assessed the incidence of CV endpoints in adults with obesity and type 2 diabetes receiving INT compared with a propensity score matched control cohort.

## METHODS

### Data sources and study design

This retrospective real-world analysis combined administrative and longitudinal claims data from the Komodo Healthcare Map for both clinic patients (INT) and an external control group. The Komodo Healthcare Map is a nationally representative database of closed and open medical and prescription claims, all-cause mortality, and demographic data for over 300 million individuals across commercial, Medicare, and Medicaid plans (15,16).

Claims data spanned January 1, 2016, through June 27, 2025. Privacy-preserving tokenization linked eligible INT participants to the Komodo database. A sample of 2.4 million patients with T2D or obesity during the same timeframe was used to construct a matched control group.

INT participants were included if they enrolled for ≥6 months between January 1, 2016, and March 1, 2025. The first day of the registration month, or a randomly assigned pseudo-registration month for controls, served as the index date. Patients with type 1 diabetes, heart failure, stage 4+ chronic kidney disease, pregnancy, cancer, or acute psychosis were ineligible for INT and therefore were excluded from the control sample. To assess primary prevention of CV events, we excluded patients with any arrhythmia, cardiac arrest, hemorrhage, stroke, myocardial infarction, ischemic heart disease, or peripheral artery disease at baseline. Those <18 years old or lacking ≥12 months of baseline data or <6 months of follow-up (allowing ≤30-day gaps) were also excluded.

Because no identifiable data were used, Institutional Review Board review and consent were not required. All records were deidentified and compliant with U.S. privacy regulations, including the Health Insurance Portability and Accountability Act of 1996.

### Telemedicine Nutrition Program

INT participants receive remote medical care for T2D or obesity, individualized health coaching, daily biometric feedback, and peer support. Patients are guided within their personally and culturally preferred cuisines to initially reduce carbohydrates to ≤30 g/day (<10% of total calories), achieving nutritional ketosis with β-hydroxybutyrate (BHB) ≥0.3 mmol/L (target 0.5– 3.0). The protein target is 1.5 g/kg reference body weight. Dietary fat is incorporated without limits on or replacements for saturated fat. Patients are advised to consume 3–5 servings of non-starchy vegetables daily, to eat to satiety, to maintain adequate hydration and mineral intake, and to seek local care for hypertension or lipid management.

### Covariate measures

Demographics included age, sex, race, U.S. region, and payer type. Baseline drug use encompassed lipid, blood pressure, and diabetes medications. Baseline comorbidities were identified by at least one ICD-10 claim during the baseline year (Appendix 1). Medication categories were defined by at least one prescription claim (Appendix 2). Follow-up drug use was measured by the proportion of days covered (PDC).

Baseline conditions included T2D, obesity, hyperlipidemia, hypertension, CKD (stages 1–3), liver disease, and smoking. Pre-existing CV disease was defined by any arrhythmia, cardiac arrest, hemorrhage, stroke, myocardial infarction, ischemic heart disease, or peripheral artery disease during the baseline year or first 90 days after index date. Rates of CV disease were comparable whether measured using one year of baseline data or all pre-index data (Appendix 3).

### Outcome measures

Outcomes were defined by ≥1 ICD-10 diagnosis on any claim occurring after the first 90 days of program participation or index date (Appendix 1). Primary outcomes included: (1) MACE-3 (nonfatal myocardial infarction, nonfatal stroke, or all-cause death); (2) MACE-6 (MACE-3 plus hospitalization for heart failure or unstable angina, or percutaneous coronary intervention); (3) new-onset CV disease (arrhythmia, cardiac arrest, hemorrhage, heart failure, ischemic heart disease, or peripheral artery disease); and (4) onset of hypertension.

Secondary outcomes included all-cause mortality. Safety outcomes included cardiac arrhythmia, atrial fibrillation, and Long QTc syndrome. An exploratory analysis examined associations between BHB levels—a marker of ketosis—and CV events within the INT group.

### Statistical analysis

The control group was constructed using 1:1 nearest-neighbor propensity score matching (PSM). Exact matching was applied to five-year age groups, sex, race, baseline T2D and obesity, and the following drug categories: anticoagulants, antiplatelets, mineralocorticoid receptor agonists, GLP-1 receptor agonists, insulin, metformin, sulfonylureas, DPP-4 inhibitors, thiazolidinediones, RAAS inhibitors, diuretics, beta blockers, statins, calcium channel blockers, and SGLT2 inhibitors (17). The propensity score also included hypertension, hyperlipidemia, CKD stages 1– 3, smoking, U.S. region, and payer type.

Time-to-event outcomes were estimated using Cox proportional hazards models. Patients were followed until a study endpoint or end of claims coverage in an intent-to-treat framework.

Because mortality data derived from multiple non-claims sources (e.g., social security administrative data, obituaries, and other administrative files), mortality was modeled using two specifications: censorship at either mortality or end of claims coverage, and censorship at mortality or end of database access (June 27, 2025).

Three sensitivity analyses tested robustness: (1) inverse probability of treatment weighting (IPTW) using the same propensity score components; (2) inclusion of follow-up PDC for lipid, blood pressure, and diabetes medications; and (3) PSM restricted to controls with overlapping index years. Matching used the *MatchIt* and *WeightIt* R packages (18,19).

In a secondary exploratory analysis, survival models for all endpoints except new-onset CV disease were repeated for INT participants with pre-existing CV disease and IPT-weighted controls. A final exploratory model examined potential mechanisms, assessing linear trends across BHB categories (<0.3, 0.3–0.5, 0.5–1.0, ≥1.0 mmol/L) coded as ordinal variables in Cox regression. Models were adjusted for age, sex, race, region, payer type, and baseline diabetes and obesity status. Reporting followed STROBE guidelines for observational studies (20).

## RESULTS

A total of 6,091 INT patients and 258,269 controls met study eligibility criteria (Table 1). Table 2 summarizes baseline characteristics before and after matching. Prior to matching, the INT and control samples had comparable age, sex, and race distributions. The INT group included more participants from the Midwest and South regions, higher prevalence of T2D, obesity, liver disease, hypertension, and hyperlipidemia, and greater use of T2D-specific and cardioprotective medications.

**Table 1.**
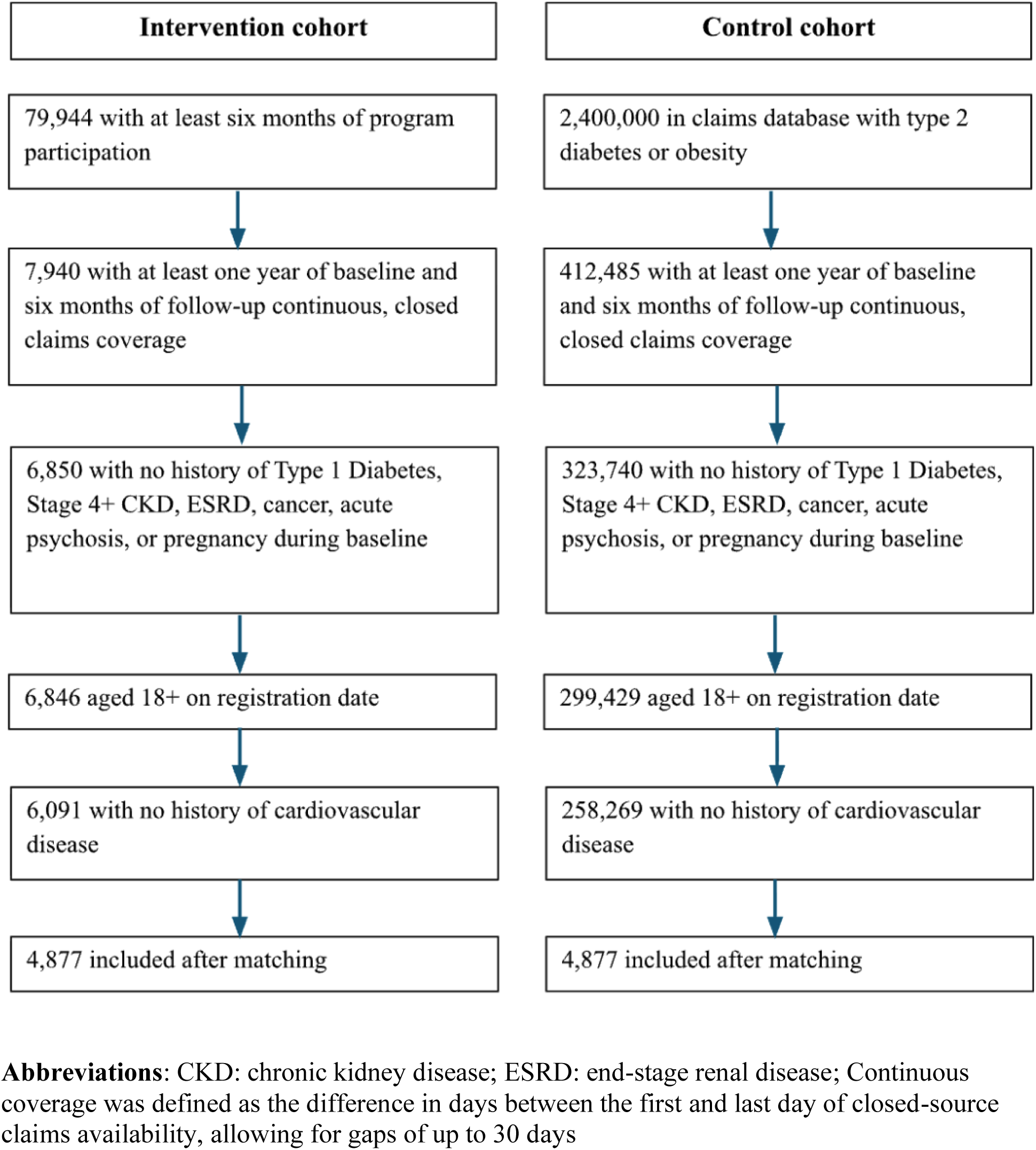
Study Sample Flowchart.

**Table 2.** Baseline Characteristics of the Study Population Before and After Propensity Score Matching.

|  | Before matching |  |  |  | After matching |  |  |  |
| --- | --- | --- | --- | --- | --- | --- | --- | --- |
| Characteristic | Intervention<br>N = 6,846 | Control<br>N = 299,429 | SMD<sup>a</sup> | p-val<sup>b</sup> | Intervention<br>N = 4,877 | Control<br>N = 4,877 | SMD | p-val |
| **Demographics** |  |  |  |  |  |  |  |  |
| Age at index date (years) | 52 (9.5) | 52 (14.9) | 0.02 | 0.44 | 51 (9.5) | 51 (9.6) | 0.00 | 0.98 |
| **Race** |  |  | 0.13 | <0.001 |  |  | 0.00 | 1.0 |
| White | 3,546 (51.8%) | 142,748 (47.7%) |  |  | 2,616 (53.6%) | 2,616 (53.6%) |  |  |
| Black or African American | 700 (10.2%) | 33,204 (11.1%) |  |  | 432 (8.9%) | 432 (8.9%) |  |  |
| Hispanic or Latino | 543 (7.9%) | 31,689 (10.6%) |  |  | 343 (7.0%) | 343 (7.0%) |  |  |
| Asian or Pacific Islander | 224 (3.3%) | 9,754 (3.3%) |  |  | 143 (2.9%) | 143 (2.9%) |  |  |
| Other | 137 (2.0%) | 9,433 (3.2%) |  |  | 69 (1.4%) | 69 (1.4%) |  |  |
| Missing | 1,696 (24.8%) | 72,601 (24.2%) |  |  | 1,274 (26.1%) | 1,274 (26.1%) |  |  |
| **Sex** |  |  | 0.06 | <0.001 |  |  | 0.00 | 1.0 |
| Female | 3,931 (57.4%) | 165,501 (55.3%) |  |  | 2,939 (60.3%) | 2,939 (60.3%) |  |  |
| Male | 2,879 (42.1%) | 131,094 (43.8%) |  |  | 1,922 (39.4%) | 1,922 (39.4%) |  |  |
| Missing | 36 (0.5%) | 2,834 (0.9%) |  |  | 16 (0.3%) | 16 (0.3%) |  |  |
| **US Region** |  |  | 0.52 |  |  |  | 0.08 | 0.003 |
| Midwest | 2,694 (39.4%) | 85,014 (28.4%) |  |  | 2,067 (42.4%) | 1,978 (40.6%) |  |  |
| Northeast | 655 (9.6%) | 82,714 (27.6%) |  |  | 420 (8.6%) | 526 (10.8%) |  |  |
| South | 2,696 (39.4%) | 86,614 (28.9%) |  |  | 1,846 (37.9%) | 1,826 (37.4%) |  |  |
| West | 797 (11.6%) | 45,023 (15.0%) |  |  | 543 (11.1%) | 547 (11.2%) |  |  |
| Unknown | 4 (0.1%) | 64 (0.0%) |  |  | 1 (0.0%) | 0 (0.0%) |  |  |
| **Payer Type** |  |  | 0.68 | <0.001 |  |  | 0.09 | <0.001 |
| Commercial | 6,424 (93.8%) | 205,293 (68.6%) |  |  | 4,654 (95.4%) | 4,560 (93.5%) |  |  |
| Medicaid | 160 (2.3%) | 29,575 (9.9%) |  |  | 104 (2.1%) | 146 (3.0%) |  |  |
| Medicare | 256 (3.7%) | 62,986 (21.0%) |  |  | 115 (2.4%) | 162 (3.3%) |  |  |
| Missing | 6 (0.1%) | 1,575 (0.5%) |  |  | 4 (0.1%) | 9 (0.2%) |  |  |
| **Baseline Comorbidities** |  |  |  |  |  |  |  |  |
| Type 2 Diabetes | 3,323 (48.5%) | 65,849 (22.0%) | 0.58 | <0.001 | 1,794 (36.8%) | 1,794 (36.8%) | 0.00 | 1.0 |
| Obesity | 4,463 (65.2%) | 163,563 (54.6%) | 0.22 | <0.001 | 3,282 (67.3%) | 3,282 (67.3%) | 0.00 | 1.0 |
| Hypertension | 3,428 (50.1%) | 133,401 (44.6%) | 0.11 | <0.001 | 1,970 (40.4%) | 2,038 (41.8%) | -0.03 | 0.16 |
| High Cholesterol | 3,611 (52.7%) | 132,509 (44.3%) | 0.17 | <0.001 | 2,182 (44.7%) | 2,221 (45.5%) | -0.02 | 0.43 |
| MASLD | 421 (6.1%) | 14,431 (4.8%) | 0.06 | <0.001 | 231 (4.7%) | 244 (5.0%) | -0.01 | 0.54 |
| CKD, Stage 1-3 | 163 (2.4%) | 10,245 (3.4%) | -0.06 | <0.001 | 72 (1.5%) | 61 (1.3%) | 0.02 | 0.34 |
| History of Smoking | 72 (1.1%) | 6,511 (2.2%) | -0.09 | <0.001 | 37 (0.8%) | 36 (0.7%) | 0.00 | 0.91 |
| **Baseline Prescription Drugs** |  |  |  |  |  |  |  |  |
| DPP4 | 326 (4.8%) | 5,994 (2.0%) | 0.15 | <0.001 | 70 (1.4%) | 70 (1.4%) | 0.00 | 1.0 |
| GLP-1 | 1,341 (19.6%) | 20,986 (7.0%) | 0.38 | <0.001 | 564 (11.6%) | 564 (11.6%) | 0.00 | 1.0 |
| Insulin | 523 (7.6%) | 10,514 (3.5%) | 0.18 | <0.001 | 139 (2.9%) | 139 (2.9%) | 0.00 | 1.0 |
| Metformin | 2,635 (38.5%) | 46,352 (15.5%) | 0.54 | <0.001 | 1,442 (29.6%) | 1,442 (29.6%) | 0.00 | 1.0 |
| SGLT2i | 739 (10.8%) | 9,540 (3.2%) | 0.30 | <0.001 | 216 (4.4%) | 216 (4.4%) | 0.00 | 1.0 |
| Sulfonylureas | 647 (9.5%) | 11,747 (3.9%) | 0.22 | <0.001 | 216 (4.4%) | 216 (4.4%) | 0.00 | 1.0 |
| Thiazolidinediones | 178 (2.6%) | 3,046 (1.0%) | 0.12 | <0.001 | 28 (0.6%) | 28 (0.6%) | 0.00 | 1.0 |
| Anticoagulants | 138 (2.0%) | 9,412 (3.1%) | -0.07 | <0.001 | 31 (0.6%) | 31 (0.6%) | 0.00 | 1.0 |
| Beta blocker | 887 (13.0%) | 43,529 (14.5%) | -0.05 | <0.001 | 281 (5.8%) | 281 (5.8%) | 0.00 | 1.0 |
| Calcium blocker | 948 (13.8%) | 41,636 (13.9%) | 0.00 | 0.89 | 419 (8.6%) | 419 (8.6%) | 0.00 | 1.0 |
| Diuretic | 1,478 (21.6%) | 58,257 (19.5%) | 0.05 | <0.001 | 799 (16.4%) | 799 (16.4%) | 0.00 | 1.0 |
| MRA | 180 (2.6%) | 5,966 (2.0%) | 0.04 | <0.001 | 60 (1.2%) | 60 (1.2%) | 0.00 | 1.0 |
| RAASi | 2,821 (41.2%) | 92,579 (30.9%) | 0.22 | <0.001 | 1,612 (33.1%) | 1,612 (33.1%) | 0.00 | 1.0 |
| Statin | 2,778 (40.6%) | 87,891 (29.4%) | 0.24 | <0.001 | 1,526 (31.3%) | 1,526 (31.3%) | 0.00 | 1.0 |

**Abbreviations:** SMD: standardized mean difference; MASLD: metabolic dysfunction-associated steatotic liver disease; CKD: chronic kidney disease; DPP-4: dipeptidyl peptidase-4; GLP-1: glucagon-like peptide-1; SGLT2i: sodium-glucose cotransporter 2 inhibitor; MRA: mineralocorticoid receptor agonist; RAASi: renin-angiotensin-aldosterone system inhibitors a SMDs were calculated as the difference in group means divided by the pooled standard deviation. b *p* values were calculated using Wilcoxon's rank-sum test for continuous variables, Fisher exact tests for binary variables, and chi-square tests for categorical variables.

After matching, the analytic sample comprised 4,877 INT participants and 4,877 matched controls. All variables achieved standardized mean differences <0.1, confirming adequate balance.

Overall, 60% of participants were female, with a mean age of 51 years at registration. Two-thirds had obesity and one-third had T2D. Fifty-four percent were White, and 95% were commercially insured. Seventy-three percent achieved biomarker levels consistent with adherence to dietary recommendations (Appendix 4).

The median (IQR) follow-up was 612 (315–820) days for INT and 806 (334–1,094) days for controls, ranging from 6 months to 8 years and 6 months to 12 years, respectively.

The incidence rate of MACE-3 was 4.1 events per 1,000 person-years in the INT group versus 9.3 in controls (hazard ratio [HR] 0.44; 95% CI 0.29–0.65; p < 0.001) across 18,957 person-years of follow-up. Findings were similar for the other primary outcome measures: for MACE-6, incidence rates were 5.7 and 10.8 per 1,000 person-years (HR 0.52; 95% CI 0.37–0.73; p < 0.001). For new-onset CV disease, rates were 27.7 and 36.9 per 1,000 person-years, with cumulative incidences of 4.65% and 8.16% (HR 0.70; 95% CI 0.59–0.82; p < 0.001). The HR for new-onset hypertension was 0.81 (95% CI 0.70–0.93; p < 0.001).

Figures 1–3 show survival curves for primary, secondary, and safety outcomes. Table 3 lists cumulative incidence and incidence per 1,000 person-years, and Appendix 5 provides full model results.

**Figure 1.**
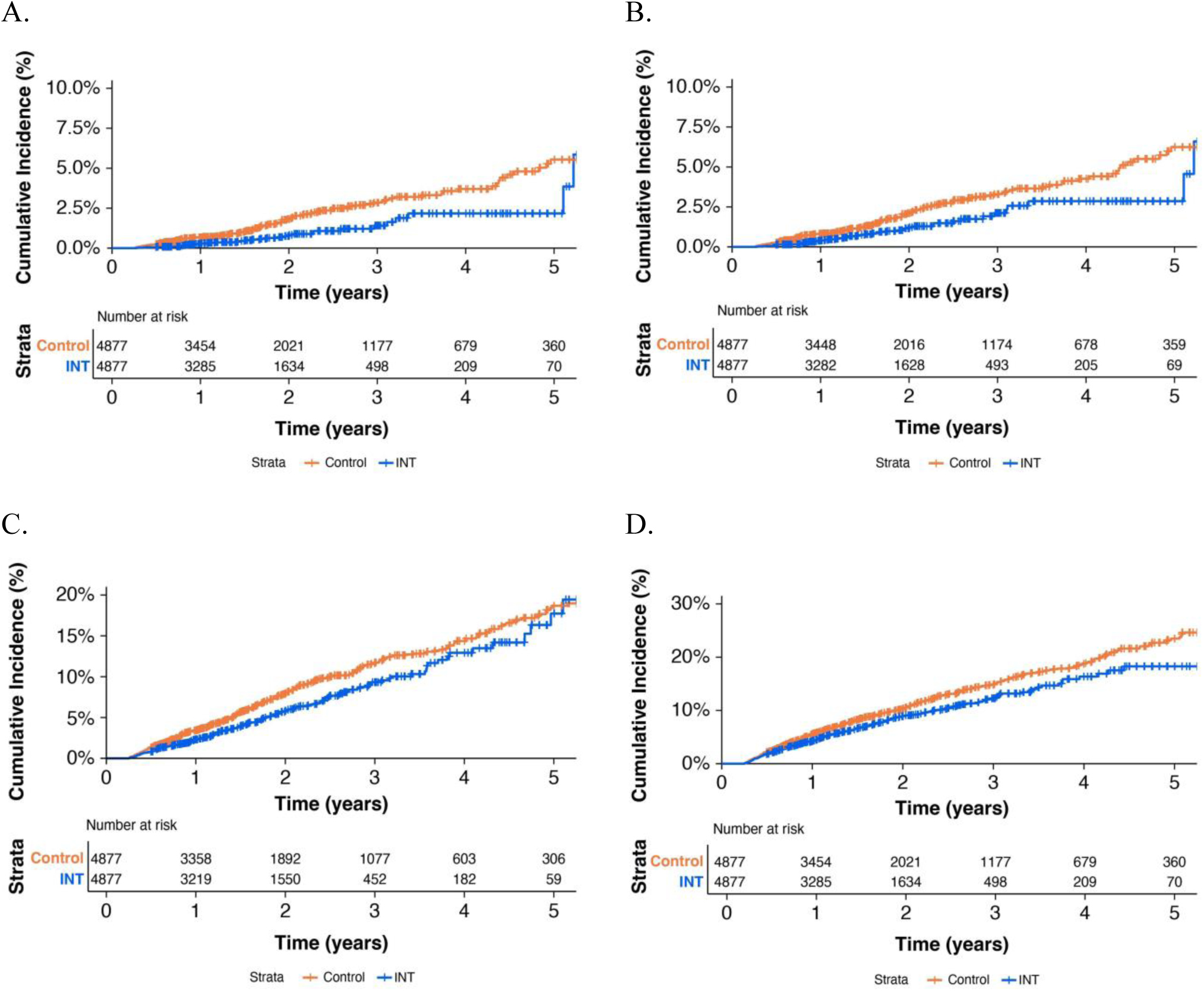
Primary outcomes. Cumulative incidence curves for (A) new-onset MACE-3, (B) new-onset MACE-6, (C) new-onset all cardiovascular disease (CVD), and (D) new-onset hypertension, comparing INT participants with matched UC controls. Curves are shown over 5 years to aid interpretation, as sample sizes decline in later follow-up. All analyses used the full observation window with standard right-censoring. At-risk tables are displayed below each panel, and steps in the curves indicate incident events. Estimates toward the end of follow-up should be interpreted cautiously due to smaller numbers at risk.

**Figure 2.**
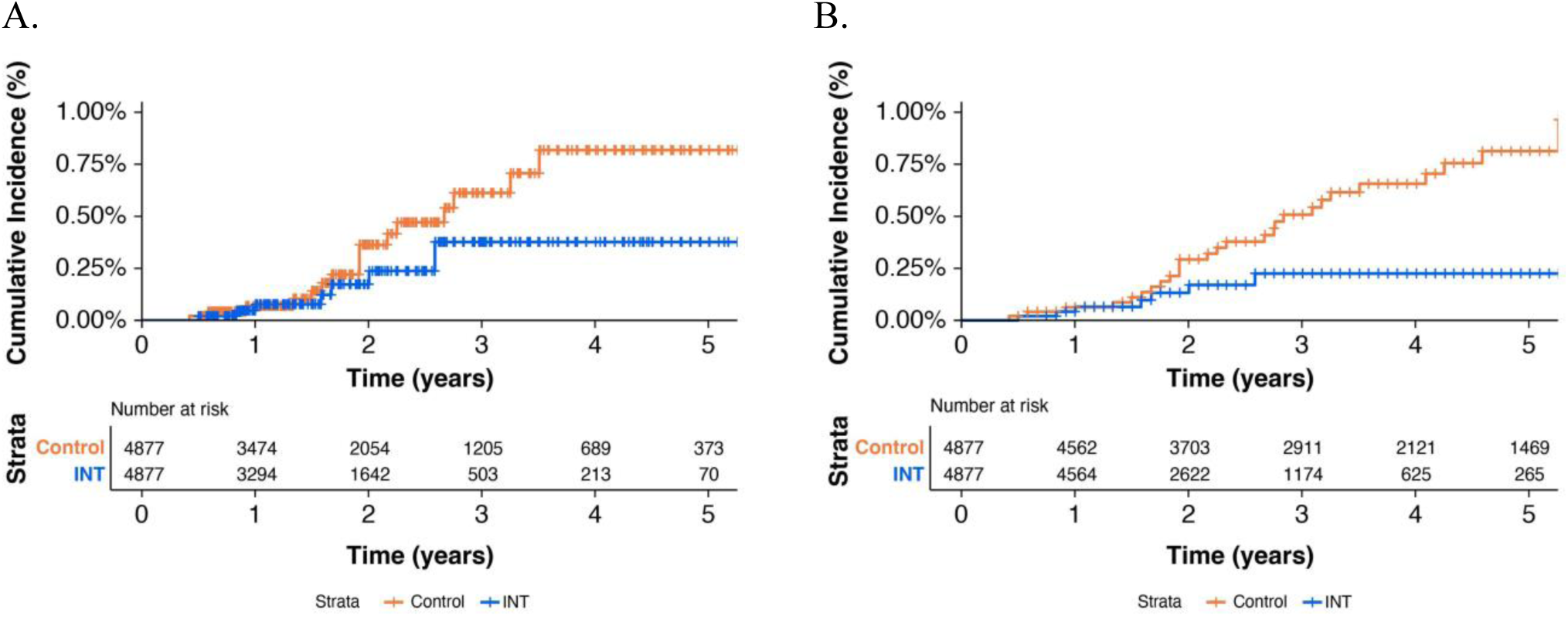
Secondary outcomes. Cumulative incidence curves for (A) mortality within the closed-claims time frame and (B) uncensored mortality, comparing INT participants with matched UC controls. Curves are displayed over 5 years to aid interpretation, as sample sizes decline during later follow-up. All analyses used the complete observation window with standard right-censoring. At-risk tables are shown below each panel, and steps in the curves represent incident events. Estimates near the end of follow-up should be interpreted with caution due to smaller numbers of participants remaining at risk.

**Figure 3.**
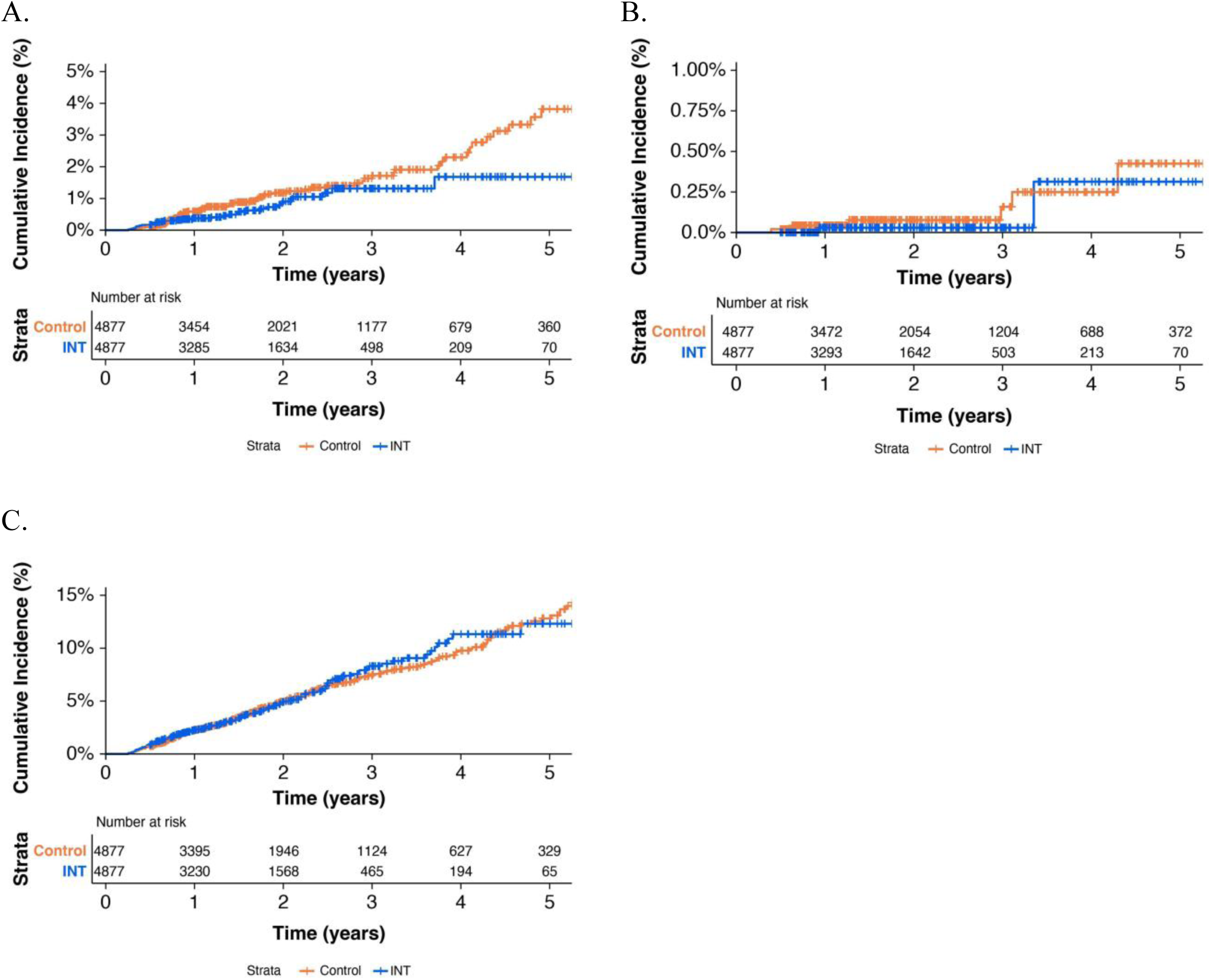
Safety outcomes. Cumulative incidence curves for (A) atrial fibrillation (B) long QT and (C) all arrhythmias, comparing INT participants with matched UC controls. Curves are shown over 5 years to aid interpretation, as sample sizes decrease during later follow-up. All analyses used the full observation window with standard right-censoring. At-risk tables are presented below each panel, and steps in the curves indicate incident events. Estimates toward the end of follow-up should be interpreted cautiously due to smaller numbers of participants remaining at risk.

**Table 3.** Incidence Rates and Hazard Ratios for Primary, Secondary, and Safety Outcomes During Follow-Up.

| Outcome | Cumulative incidence rate<sup>a</sup> |  | Incidence per 1,000 person-years<sup>b</sup> |  | Cox model estimates<sup>c</sup> |  |  |
| --- | --- | --- | --- | --- | --- | --- | --- |
|  | Control | INT | Control | INT | HR | 95% CI | *p*-value |
| **Primary outcomes** |  |  |  |  |  |  |  |
| MACE-3 | 2.1% | 0.7% | 9.3 | 4.1 | 0.44 | (0.29, 0.65) | <0.001 |
| MACE-6 | 2.4% | 1.0% | 10.8 | 5.7 | 0.52 | (0.37, 0.73) | <0.001 |
| All CV Disease | 8.2% | 4.7% | 36.9 | 27.7 | 0.70 | (0.59, 0.82) | <0.001 |
| Hypertension | 10.9% | 7.0% | 49.3 | 41.8 | 0.81 | (0.70, 0.93) | <0.001 |
| **Secondary outcomes** |  |  |  |  |  |  |  |
| Death | 0.4% | 0.1% | 1.6 | 0.8 | 0.47 | (0.19, 1.14) | 0.10 |
| Death, Uncensored | 0.8% | 0.1% | 3.7 | 0.8 | 0.36 | (0.16, 0.84) | 0.02 |
| **Safety outcomes** |  |  |  |  |  |  |  |
| Atrial Fibrillation | 1.4% | 0.7% | 6.3 | 4.1 | 0.65 | (0.42, 1.00) | 0.05 |
| All Arrhythmias | 5.4% | 4.1% | 24.5 | 24.2 | 1.00 | (0.83, 1.21) | 0.97 |
| Long QT Syndrome | 0.1% | 0.0% | 0.6 | 0.2 | 0.46 | (0.09, 2.33) | 0.35 |
| Mean follow-up, days | 806 | 612 |  |  |  |  |  |
| Total follow-up, person-years | 10,769 | 8,177 |  |  |  |  |  |

**Abbreviations:** MACE-3: major adverse cardiac events, three-point composite endpoint; MACE-6: major adverse cardiac events, six-point composite endpoint; CV: cardiovascular; HR: hazard ratio; CI: confidence interval.
a Cumulative incidence is calculated as the total number of persons experiencing the specified endpoint divided by the total number of persons.
b Incidence rates are calculated as the total number of persons experiencing the specified endpoint divided by the total person-time at risk and are expressed per 1,000 person-years.
c Hazard ratios, 95% CIs and *p*-values were estimated using Cox proportional hazards regression models adjusted for age, sex, and baseline covariates.

All-cause mortality was significantly lower across the full follow-up period (HR 0.36 for the INT group; 95% CI 0.16–0.84; p = 0.02), but not during the closed-claims period (HR 0.47; 95% CI 0.19–1.14).

For safety outcomes, HRs were not statistically significant and numerically less than 1 for new-onset atrial fibrillation (HR = 0.65; 95% CI 0.42–1.01) and arrhythmia (HR = 1.00; 95% CI 0.83–1.21). Ten Long QTc events occurred overall (HR = 0.46; 95% CI 0.09–2.33).

Table 4 summarizes sensitivity analyses. The IPTW-ATT model produced slightly smaller but directionally consistent results. The PSM model adjusted for cardioprotective medication use (PDC) yielded slightly stronger associations, indicating reduced medication reliance in INT (Appendix 6). The PSM model restricted to controls with overlapping index years showed directionally consistent but statistically insignificant impacts on the onset of hypertension, smaller but significant impacts on the onset of MACE-6, MACE-3, hypertension, and death during the entire follow-up period.

**Table 4.** Sensitivity Analyses Using Alternative Modeling Approaches for Primary, Secondary, and Safety Outcomes.

|  | IPTW |  |  | PSM adj. for PDC |  |  | PSM incl. registration year |  |  |
| --- | --- | --- | --- | --- | --- | --- | --- | --- | --- |
| Outcome | HR<sup>a</sup> | 95% CI | *p*-value | HR | 95% CI | *p*-value | HR | 95% CI | *p*-value |
| *Primary outcomes* |  |  |  |  |  |  |  |  |  |
| MACE-3 | 0.51 | (0.39, 0.67) | <0.001 | 0.44 | (0.29, 0.65) | <0.001 | 0.57 | (0.41, 0.80) | <0.001 |
| MACE-6 | 0.57 | (0.46, 0.72) | <0.001 | 0.50 | (0.35, 0.71) | <0.001 | 0.62 | (0.46, 0.83) | <0.001 |
| All CV Disease | 0.76 | (0.68, 0.85) | <0.001 | 0.69 | (0.58, 0.82) | <0.001 | 0.71 | (0.61, 0.82) | <0.001 |
| Hypertension | 0.79 | (0.72, 0.87) | <0.001 | 0.82 | (0.71, 0.94) | 0.01 | 0.89 | (0.78, 1.01) | 0.08 |
| *Secondary outcomes* |  |  |  |  |  |  |  |  |  |
| Death | 0.54 | (0.30, 0.96) | 0.04 | 0.50 | (0.20, 1.21) | 0.12 | 0.63 | (0.29, 1.37) | 0.24 |
| Death, Uncensored | 0.38 | (0.22, 0.67) | <0.001 | 0.38 | (0.16, 0.86) | 0.02 | 0.41 | (0.21, 0.80) | 0.01 |
| *Safety outcomes* |  |  |  |  |  |  |  |  |  |
| Atrial Fibrillation | 0.92 | (0.63, 1.33) | 0.65 | 0.66 | (0.43, 1.01) | 0.05 | 0.92 | (0.63, 1.33) | 0.65 |
| All Arrhythmias | 1.04 | (0.87, 1.24) | 0.67 | 0.99 | (0.82, 1.20) | 0.91 | 1.04 | (0.87, 1.24) | 0.67 |
| Long QT Syndrome | 0.35 | (0.07, 1.75) | 0.20 | 0.53 | (0.10, 2.74) | 0.45 | 0.35 | (0.07, 1.75) | 0.20 |

**Abbreviations:** IPTW: inverse probability of treatment weighting; PSM: propensity score matching; PDC: proportion of days covered; MACE-3: major adverse cardiac events, three-point composite endpoint; MACE-6: major adverse cardiac events, six-point composite endpoint; CV: cardiovascular; HR: hazard ratio; CI: confidence interval.
<sup>a</sup> Hazard ratio, 95% CIs and *p*-values were estimated using Cox proportional hazards regression models adjusted for age, sex, and baseline covariates.
**Note:** Hazard ratios, 95% confidence intervals (CIs), and *p*-values were estimated using Cox proportional hazards regression models adjusted for age, sex, and baseline covariates. Models were conducted using three alternative approaches: (1) IPTW to balance baseline characteristics across cohorts, (2) PSM adjusted for medication adherence (PDC), and (3) PSM including registration year as an additional covariate.

The secondary exploratory analysis assessed 613 INT participants with baseline CV disease and 29,969 IPT-weighted matched controls. No statistically significant effects were observed for primary, secondary, or safety endpoints (Appendix 7).

In an exploratory analysis of dietary adherence, BHB levels modeled as ordinal categories (<0.3, 0.3–0.5, 0.5–1.0, ≥ 1.0 mmol/L) were inversely associated with new-onset CV disease (HR = 0.86; 95% CI 0.77–0.96; p = 0.01), indicating a 14% lower hazard per category increase in ketosis. Ketosis level was not significantly associated with MACE-3, MACE-6, or hypertension (all p > 0.05), possibly reflecting limited power for these less frequent outcomes (Appendix 8).

## DISCUSSION

In this claims-based analysis, patients in the intervention group were at reduced risk of cardiovascular events compared to those in a propensity score matched control group. Reductions were observed across all major outcomes, including 3-point MACE (56% reduction), 6-point MACE (48% reduction), CV events (30% reduction), new-onset hypertension (19% reduction), and all-cause mortality (53–64% reduction, directionally consistent though not significant). There was no increase in arrhythmias or long QT, and subgroup analyses revealed no excess risk among participants with prior CV disease. To our knowledge, this is the first claims-based evaluation of a nutrition intervention and the first report of CV event rates among individuals counseled on ketogenic nutrition.

### Cardiovascular Events in other Nutrition Interventions

These findings align with prior nutrition trials demonstrating that dietary interventions can meaningfully reduce CV risk. Mediterranean diet trials, including the Lyon Diet Heart Study (21), CORDIOPREV (22), and PREDIMED, have shown benefits for both secondary and primary prevention, with PREDIMED reporting a 35% reduction in primary CV events over five years (23). In contrast, other lifestyle interventions have yielded less consistent results. The Look AHEAD trial, for example, found no overall reduction in CV events after a decade of intensive calorie reduction and exercise (24), with benefit limited to participants achieving the greatest weight loss and fitness gains (25). Diabetes prevention interventions have successfully delayed diabetes onset but show limited or delayed impact on CV outcomes, with benefits emerging only after decades in the Da Qing Diabetes Prevention Outcome Study (26, 27) or not at all in the Diabetes Prevention Program Outcomes Study (28).

### Mediators of Cardiovascular Risk Reduction with Carbohydrate Reduced Nutrition Counseling

Against this backdrop, carbohydrate-reduced and ketogenic nutrition have emerged as increasingly utilized interventions targeting key cardiovascular risk factors such as T2D and obesity. While these diets share some features with Mediterranean patterns, they diverge by sharply limiting fruits, whole grains, and legumes, and are typically higher in total and saturated fat. Despite these differences, a mounting body of evidence demonstrates broad cardiometabolic benefits of ketogenic interventions, including improvements in weight, hemoglobin A1c, glycemic variability, insulin resistance, visceral fat, hepatic steatosis, blood pressure, LDL subfraction, HDL-cholesterol, triglycerides, inflammation, kidney disease progression, medication requirements (6, 9, 11, 29–38), and sustained diabetes remission (39). Collectively, these improvements in residual CV risk factors that are features of the cardiovascular-kidney-metabolic syndrome (40) support the hypothesis that ketogenic nutrition confers cardioprotection (41), a relationship corroborated by the robust associations observed in this claims-based analysis.

There was an association between increasing level of ketosis and new onset of CV disease pointing toward a likely causal effect of ketosis on CV outcomes. Consistent with this, in a separate claims analysis, on patients from the same clinic with mean BHB levels ≥0.3 mmol/L during the first six months of care were less likely to develop new-onset chronic kidney disease (CKD) than those with lower BHB levels (42). Taken together, the impact of the intervention on comprehensive residual risk markers including CKD risk potentially explains the long term protection from CV disease endpoints.

Concerns regarding long-term safety, particularly related to saturated fat intake and potential increases in LDL-C have constrained prospective investigation of the impact on CV event rates and adoption of ketogenic dietary patterns (43–45). LDL-C elevations are common, but not universal, may be transient (39, 46), and may occur alongside shifts toward less atherogenic lipoprotein subtypes (11, 47, 48). At the same time, even with optimally managed LDL-C, residual CV risk remains substantial (3, 49), and the wide-ranging improvements associated with ketogenic nutrition are hypothesized to exert protective effects (41). Nonetheless, LDL-C remains a central therapeutic target in CV prevention (50), and interventions that may increase LDL-C deserve scrutiny. The present study observed an overall reduction in cardiovascular events across the intervention cohort, suggesting that the broader cardio-kidney-metabolic effects mitigate residual cardiovascular risk and may outweigh the potential impact of isolated LDL-C changes. These findings also support the rationale for prospective evaluation of cardioprotective potential in randomized clinical trials.

Sustainability is another common concern given the challenges of long-term adherence to any lifestyle change. Continuous remote care including biomarker monitoring may mitigate these barriers by providing frequent contact and real-time, individualized support. As an intent-to-treat study, this analysis provides an estimate of INT’s CV impact in a real-world setting. The observed CV event reductions are unlikely to be explained by greater medication adherence due to more frequent care team contact, as prescription claims for several cardioprotective medications were lower in the treated cohort.

## Strengths and Limitations

This study is the first to leverage claims data to evaluate CV outcomes associated with a nutrition intervention implemented in a pragmatic, real-world setting. Prior investigations of ketogenic nutrition interventions have examined changes in CV risk factors rather than CV outcomes.

Distinguishing this study from most dietary investigations, the inclusion of ketosis level data in the intervention group provides objective evidence of dietary adherence, and the ability to explore mechanisms. Accordingly, this claims-based analysis addresses a critical gap in the literature by evaluating a spectrum of hard CV endpoints associated with a carbohydrate-reduced and ketogenic nutrition intervention.

Several limitations must be acknowledged. First, although comprehensive matching and weighting methods were used to minimize confounding between the intervention and control groups, the observational study design cannot eliminate potential bias from unmeasured confounders. Second, claims data may undercapture diagnoses and procedures used to construct both covariates and outcomes during one-year baseline and follow-up periods, leading to misclassification or inaccurate measurement of incidence. Both the intervention and control arms had continuous claims coverage and were matched closely on baseline engagement in care, likely limiting the impact of undercaptured diagnoses. Third, because nearly 90% of participants had commercial insurance, additional evidence is needed to determine the generalizability of these findings to non–commercially insured populations. Fourth, because the study did not involve linked electronic health record data, the relative contributions of changes in specific risk factors such as weight, glycemia, insulin resistance, central abdominal fat, blood pressure, inflammation, and LDL-C is beyond the scope of this analysis.

Despite these limitations, the consistent association between ketogenic nutrition-oriented care and lower incidence of CV events provides a strong signal for the safety and cardioprotective nature of the intervention.

## CONCLUSIONS

Our findings suggest that a telemedicine program emphasizing carbohydrate-reduced nutrition exerts cardioprotective effects as evidenced by lower CV event rates among participants.

## Data Availability

The data underlying this article was provided by the third party, Komodo Health, under license and cannot be shared publicly. The source data for this study were licensed by Virta Health from Komodo Health, and hence may not be shared publicly.

## ACKNOWLEDGMENTS

PVS and CGPR contributed equally as co-first authors. Manuscript drafting was conducted by PVS and CGPR. All authors provided critical review and final approval. PVS conducted the data analysis. PVS, SJA, and FLJ had access to the data.

## Substantial contributions, non-author

Rebecca N. Adams, PhD reviewed the manuscript.

