## Appendix for "Real-World Cardiovascular Outcomes with a Carbohydrate-Reduced Telemedicine Intervention"

**Appendix 1.** ICD-10 Diagnosis code categories

| ICD-10 | Description |
|---|---|
| Stroke (I63.*) | |
| I63.* | Cerebral infarction |
| Hemorrhage (I60.*, I61.*, I62.*) | |
| I60.00-9 | Subarachnoid hemorrhage |
| | Subcortical hemorrhage |
| | Subarachnoid hemorrhage from carotid siphon and bifurcation |
| | Intracerebral hemorrhage, multiple localized |
| | Subarachnoid hemorrhage from vertebral artery |
| I62 | Non-traumatic subdural hemorrhage |
| | Non-traumatic intracerebral ventricular hemorrhage |
| | Spontaneous subarachnoid hemorrhage |
| I61.X | Cortical hemorrhage |
| | Brain stem hemorrhage |
| | Spontaneous hemorrhage of cerebral hemisphere |
| | Spontaneous cerebellar hemorrhage |
| | Spontaneous cerebral hemorrhage |
| Arrhythmias (I47-49, R00.1) | |
| I48.* | Atrial fibrillation |
| I47.*, I49.1, R00.1 | Paroxysmal tachycardia |
| | Atrial flutter |
| Ischemic Heart Disease (I20, I23, I24, I25) | |
| I24 | Acute ischemic heart disease |
| Myocardial Infarction (I21) | |
| I21.X | Acute non-ST segment elevation myocardial infarction |
| | Acute ST segment elevation myocardial infarction |
| | Myocardial infarction |
| | Myocardial infarction due to demand ischemia |
| | Acute ST segment elevation myocardial infarction involving left anterior descending coronary artery |
| | Acute ST segment elevation myocardial infarction due to left coronary artery occlusion |
| | Acute ST segment elevation myocardial infarction due to right coronary artery occlusion |
| Heart failure (I50, I13, I11) | |
| I50.X, I13.0, I11.0 | Congestive heart failure |
| | Hypertensive heart and renal disease with (congestive) heart failure |
| | Left heart failure |
| | Systolic heart failure |
| | Diastolic heart failure |

| | |
|---|---|
| | Hypertensive heart failure |
| | High output heart failure |
| | Chronic right-sided heart failure |
| | Right heart failure secondary to left heart failure |
| | Chronic congestive heart failure |
| | Biventricular congestive heart failure |
| | Acute on chronic right-sided congestive heart failure |
| | Chronic systolic heart failure |
| | Chronic diastolic heart failure |
| | Acute on chronic systolic heart failure |
| | Acute on chronic diastolic heart failure |
| | Chronic combined systolic and diastolic heart failure |
| | Acute on chronic combined systolic and diastolic heart failure |
| | Acute right-sided heart failure |
| | Acute systolic heart failure |
| | Acute diastolic heart failure |
| | Acute combined systolic and diastolic heart failure |
| | Heart Failure |
| Cardiac arrest | |
| I46.9 | Cardiac arrest |
| I46.2 | Cardiac arrest due to cardiac disorder |
| I97.* | Cardiac arrest as a complication of care |
| R57.0 | Cardiogenic shock |
| Peripheral Arterial Diseases | |
| I70.*, I77.7* | Peripheral arterial diseases |
| Hypertension | |
| I10 | Essential |
| I11 | Hypertensive heart disease |
| I12 | Hypertensive chronic kidney disease |
| I13 | Hypertensive heart & chronic kidney disease |
| I15.0,2,9 | Secondary hypertension |
| I16.0,1 | Hypertensive crisis |
| Other baseline comorbidities | |
| E11.* | Diabetes (Type 2) |
| E10.* | Diabetes (Type 1) |
| E66.*, Z68.25-29, Z68.3*, Z68.4* | Obesity or overweight or BMI 25+ |
| N181.*-N186.* | CKD Stage 1 - ESRD |
| I4581 | Long QTc |
| O* or Z3* | Pregnancy |
| E78 | High Cholesterol |
| Z720.* | Smoking |
| C* | Cancer |
| Acute psychosis | |

| | |
|---|---|
| F20 | Schizophrenia |
| F21 | Schizotypal disorder |
| F22 | Delusional disorders |
| F23 | Brief psychotic disorders |
| F24 | Shared psychotic disorder |
| F25 | Schizoaffective disorder |
| F28 | Other psychotic disorder not due to a substance or known psychological condition |
| F29 | Unspecified psychosis not due to a substance or known psychological condition |

**Appendix 2.** Generic Drug Names and Prescription Drug Categories

| Drug category | Generic names |
|---|---|
| SGLT2 | Canagliflozin<br>Dapagliflozin<br>Empagliflozin<br>Ertugliflozin |
| Sulfonylureas | Glipizide<br>Glyburide<br>Glimepiride<br>Chorpropamide<br>Tolbutamide<br>Tolazamide<br>Acetohexamide |
| DPP4 | Sitagliptin<br>Saxagliptin<br>Linagliptin<br>Alogliptin<br>Vildagliptin<br>Teneligliptin |
| Thiazoledinedione | Pioglitazone<br>Rosiglitazone |
| Insulin | Insulin<br>Humulin<br>Humalog<br>Novolog |
| GLP-1 | Exenatide<br>Liraglutide<br>Lixisenatide<br>Dulaglutide<br>Semaglutide<br>Tirzepatide |
| Anticoagulant | Warfarin<br>Dabigatran<br>Rivaroxaban<br>Apixaban<br>Edoxaban<br>Betrixaban |
| Antiplatelets | Anagrelide<br>Colostazol<br>Clopidogrel<br>Dipyridamole<br>Prasugrel<br>Icagrelor<br>Ticlopidine<br>Vorapaxar |

| | |
|---|---|
| | Cangrelor |
| Statins | Atorvastatin<br>Simvastatin<br>Rosuvastatin<br>Pravastatin<br>Lovastatin<br>Fluvastatin<br>Pitavastatin<br>Ezetimibe<br>Alirocumab<br>Inclisiran |
| MRA | Eplerenone<br>Spironolactone |
| Diuretic | Bendroflumethiazide<br>Chlorothiazide<br>Chlorthalidone<br>Hydrochlorothiazide<br>Indapamide<br>Myethoclothiazide<br>Metolazone<br>Bumetanide<br>Ethacrynate sodium<br>Ethacrynic acid<br>Furosemide<br>Torsemide<br>Amiloride<br>Triamterene |
| RAAS Inhibitors | Benazepril<br>Captopril<br>Enalapril<br>Fosinopril<br>Lisinopril<br>Moexipril<br>Perindopril<br>Quinapril<br>Ramipril<br>Trandolapril<br>Azilsartan<br>Candesartan<br>Eprosartan<br>Irbesartan<br>Losartan<br>Olmesartan<br>Telmisartan<br>Valsartan<br>Aliskiren |

| | |
|---|---|
| Beta Blockers | Atenolol<br>Betaxolol<br>Bisoprolol<br>Metoprolol tartrate<br>Metoprolol succinate<br>Nebivolol<br>Nadolol<br>Propanolol<br>Acebutolol<br>Pindolol<br>Timolol<br>Carvedilol<br>Labetalol<br>Esmolol<br>Sotalol |
| Calcium Blockers | Amlodipine<br>Felodipine<br>Isradipine<br>Nicardipine<br>Nifedipine<br>Nisoldipine<br>Clevidipine<br>Nimodipine<br>Diltiazem<br>Verapamil |

**Appendix 3.** Rates of Baseline CVD outside the 1-year pre-index/enrollment closed claims period

| Baseline CVD using all historical claims | Matched Control N = 4,877 | INT N = 4,877 | p-value |
|---|---|---|---|
| Atrial Fibrillation | 38 (0.8%) | 28 (0.6%) | 0.2 |
| Arrhythmia | 214 (4.4%) | 314 (6.4%) | <0.001 |
| Cardiac Arrest | 6 (0.1%) | 8 (0.2%) | 0.6 |
| Myocardial Infarction | 17 (0.3%) | 24 (0.5%) | 0.3 |
| Hemorrhage | 8 (0.2%) | 5 (0.1%) | 0.4 |
| Heart Failure | 75 (1.5%) | 78 (1.6%) | 0.8 |
| Ischemic Heart Disease | 152 (3.1%) | 165 (3.4%) | 0.5 |
| Peripheral Artery Disease | 31 (0.6%) | 44 (0.9%) | 0.13 |
| Stroke | 23 (0.5%) | 19 (0.4%) | 0.5 |
| Any CVD | 466 (9.6%) | 558 (11%) | 0.002 |

**Note**: Baseline cardiovascular disease (CVD) events were identified using all available historical claims preceding the 1-year pre-index/enrollment closed-claims period. Differences between intervention (INT) and matched control cohorts were assessed using chi-square tests for categorical variables.

**Abbreviations:** CVD, cardiovascular disease; INT, individualized nutrition therapy

**Appendix 4**. Program participation measures for INT cohort

| **Program Participation Measures** | **INT**<br>**N = 4,877** |
|---|---|
| *Starting HbA1c* | |
| <6.5 | 2,815 (57.7%) |
| 6.5-7.0 | 428 (8.8%) |
| 7.0-8.0 | 427 (8.8%) |
| 8.0-9.0 | 219 (4.5%) |
| >9.0 | 305 (6.3%) |
| Missing | 683 (14.0%) |
| *Starting BMI* | |
| <18.5 | 0 (0.0%) |
| 18.5-24.9 | 123 (2.5%) |
| 25.0-29.9 | 621 (12.7%) |
| 30.0-34.9 | 1,670 (34.2%) |
| 35.0-39.9 | 1,276 (26.2%) |
| >40.0 | 1,186 (24.3%) |
| Missing | 1 (0.0%) |
| *Ketone levels in the first 180 days* | |
| 0 | 191 (3.9%) |
| 0-0.3 | 1,149 (23.6%) |
| 0.3-0.5 | 1,479 (30.3%) |
| 0.5-1.0 | 1,602 (32.8%) |
| ≥1.0 | 456 (9.4%) |
| *Program participation duration* | |
| 6 to 12 months | 1,881 (38.6%) |
| 12 to 24 months | 1,826 (37.4%) |
| ≥24 months | 1,170 (24.0%) |

**Note:** Program participation metrics were derived from participants enrolled in the intervention (INT) program. Starting HbA1c and BMI values represent measurements at program initiation. Ketone levels reflect the mean beta-hydroxybutyrate (BHB) concentrations recorded during the first 180 days of participation. Program participation duration corresponds to the total period of continuous engagement in the intervention. Percentages are based on non-missing data.
**Abbreviations:** HbA1c, hemoglobin A1c; BMI, body mass index; BHB, beta-hydroxybutyrate; INT, intervention.

**Appendix 5.** Multivariable Cox Regression Models for Primary Cardiovascular Outcomes in the Matched Cohort

| | **MACE 3** | | **MACE 6** | | **All CVD** | | **Hypertension** | |
|---|---|---|---|---|---|---|---|---|
| **Variables** | Hazard Ratio (95% CI) | p | Hazard Ratio (95% CI) | p | Hazard Ratio (95% CI) | p | Hazard Ratio (95% CI) | p |
| INT | 0.44 (0.29, 0.65) | <0.001 | 0.52 (0.37, 0.73) | <0.001 | 0.7 (0.59, 0.82) | <0.001 | 0.81 (0.70, 0.93) | <0.001 |
| Payer Type: Commercial | 4183897.92 | 1 | 1780434.62 | 0.99 | 1.34 (0.19, 9.59) | 0.77 | 1.81 (0.25, 12.97) | 0.55 |
| Payer Type: Medicaid | 6683452.46 | 1 | 3420067.98 | 0.99 | 2.27 (0.30, 16.97) | 0.43 | 1.64 (0.22, 12.29) | 0.63 |
| Payer Type: Medicare | 3877368.64 | 1 | 1927046.32 | 0.99 | 1.90 (0.26, 13.90) | 0.53 | 2.08 (0.28, 15.34) | 0.47 |
| US Region: Northeast | 1.22 (0.72, 2.07) | 0.47 | 0.99 (0.59, 1.65) | 0.96 | 1.16 (0.89, 1.51) | 0.28 | 1.11 (0.89, 1.40) | 0.35 |
| US Region: South | 1.11 (0.74, 1.65) | 0.62 | 1.16 (0.81, 1.68) | 0.42 | 1.38 (1.14, 1.66) | <0.001 | 1.12 (0.96, 1.31) | 0.15 |
| US Region: West | 0.66 (0.34, 1.29) | 0.22 | 1.04 (0.62, 1.73) | 0.89 | 1.11 (0.85, 1.45) | 0.43 | 1.05 (0.84, 1.31) | 0.68 |
| Age at index date | 1.05 (1.03, 1.08) | <0.001 | 1.05 (1.03, 1.07) | <0.001 | 1.06 (1.05, 1.07) | <0.001 | 1.00 (0.99, 1.01) | 0.52 |
| Female | Referent | 1 | Referent | 1 | Referent | 1 | Referent | 1 |
| Male | 1.28 (0.90, 1.83) | 0.17 | 1.34 (0.97, 1.84) | 0.07 | 1.38 (1.17, 1.63) | <0.001 | 1.04 (0.91, 1.20) | 0.55 |
| White | Referent | 1 | Referent | 1 | Referent | 1 | Referent | 1 |
| Black or African American | 1.32 (0.72, 2.43) | 0.37 | 1.12 (0.63, 1.99) | 0.71 | 0.89 (0.65, 1.21) | 0.45 | 1.05 (0.82, 1.34) | 0.70 |
| Hispanic or Latino | 0.98 (0.44, 2.15) | 0.95 | 1.03 (0.53, 2.01) | 0.93 | 1.09 (0.79, 1.51) | 0.6 | 1.06 (0.82, 1.39) | 0.65 |
| Asian or Pacific Islander | 0.58 (0.14, 2.39) | 0.45 | 0.64 (0.20, 2.05) | 0.46 | 1.02 (0.64, 1.63) | 0.92 | 1.10 (0.77, 1.58) | 0.59 |
| Other Race | 2.18 (0.68, 6.99) | 0.19 | 1.66 (0.52, 5.27) | 0.39 | 1.71 (0.96, 3.05) | 0.07 | 1.08 (0.61,1.91) | 0.81 |
| Missing Race | 1.22 (0.81, 1.84) | 0.35 | 1.09 (0.75, 1.59) | 0.66 | 0.94 (0.77, 1.15) | 0.57 | 0.92 (0.78, 1.08) | 0.31 |
| Baseline Obesity | 1.04 (0.73, 1.49) | 0.84 | 0.99 (0.72, 1.37) | 0.97 | 1.02 (0.86, 1.20) | 0.81 | 0.64 (0.56, 0.74) | <0.001 |
| Baseline Type 2 diabetes | 1.45 (1.01, 2.09) | 0.05 | 1.68 (1.20, 2.24) | 0 | 1.34 (1.14, 1.59) | <0.001 | 0.79 (0.68, 0.92) | <0.001 |

**Note:** Hazard ratios (HRs), 95% confidence intervals (CIs), and *p*-values were estimated using Cox proportional hazards models adjusted for age, sex, race or ethnicity, payer type, U.S. region, baseline obesity, and baseline type 2 diabetes. Models were stratified by matched set and represent fully adjusted results for the matched intervention and control cohorts. Hazard ratio estimates for covariates with very low event counts (e.g., certain payer categories) may be unstable due to sparse data and should be interpreted with caution.
**Abbreviations:** MACE-3, major adverse cardiac events (three-point composite endpoint); MACE-6, major adverse cardiac events (six-point composite endpoint); CVD, cardiovascular disease; HR, hazard ratio; CI, confidence interval; INT, intervention; PDC, proportion of days covered.

**Appendix 6.** Post-Intervention Differences in Cardiometabolic Medication Utilization Between INT and Control Cohorts

| | **Differences-in-Differences Model** | | | **p-value, parallel baseline trends** |
|---|---|---|---|---|
| | **Treatment Effect** | **SE** | **p-val** | |
| ***Cardiometabolic medication use, $ PMPM*** | | | | |
| **Beta blockers** | 0 | 0.09 | 0.476 | 0.557 |
| **Calcium blockers** | 0 | 0.05 | 0.313 | 0.064 |
| **Diuretics** | −0.21 | 0.09 | 0.021 | 0.122 |
| **DPP4s** | −2.42 | 0.75 | 0.001 | 0.145 |
| **GLP-1s** | −4.00 | 4.30 | 0.353 | 0.002 |
| **Insulin** | −4.51 | 1.30 | 0.001 | 0.817 |
| **Metformin** | −2.35 | 0.76 | 0.002 | 0.082 |
| **RAAS inhibitors** | −0.04 | 0.15 | 0.784 | 0.297 |
| **SGLT2i** | −11.81 | 1.75 | 0.000 | 0.964 |
| **Statin** | 0.16 | 0.21 | 0.448 | 0.933 |
| **Sulfonylureas** | −0.12 | 0.02 | 0.000 | 0.209 |
| **Thiazolidinediones** | −0.04 | 0.01 | 0.002 | 0.542 |

**Note:** Results are derived from a differences-in-differences (DID) model comparing changes in cardiometabolic medication spending between the intervention (INT) and control cohorts following program initiation. Treatment effects represent post-intervention differences adjusted for parallel baseline trends.
**Abbreviations:** DPP4s, dipeptidyl peptidase-4 inhibitors; GLP-1s, glucagon-like peptide-1 receptor agonists; RAAS, renin–angiotensin–aldosterone system; SGLT2i, sodium–glucose cotransporter-2 inhibitors; SE, standard error; PMPM, per member per month; PDC, proportion of days covered; INT, intervention.

**Appendix 7.** Survival Analyses of New-Onset Secondary Cardiovascular Events Among Participants with Existing CVD at Baseline (IPTW Model)

| **Outcome** | **Control** | **INT** | **HR** | **CI** | **p-value** |
|---|---|---|---|---|---|
| *Primary outcomes* | | | | | |
| MACE-3 | 0.0832 | 0.0503 | 1.20 | (0.87, 1.65) | 0.27 |
| MACE-6 | 0.1015 | 0.0675 | 1.18 | (0.90, 1.56) | 0.23 |
| *Secondary outcomes* | | | | | |
| Death | 0.0275 | 0.0026 | 0.37 | (0.09, 1.47) | 0.16 |
| Death (uncensored) | 0.0483 | 0.0040 | 0.32 | (0.10, 0.99) | 0.05 |
| *Safety outcomes* | | | | | |
| Atrial Fibrillation | 0.0482 | 0.0278 | 1.04 | (0.67, 1.61) | 0.87 |
| LongQT | 0.0060 | 0.0040 | 1.06 | (0.34, 3.32) | 0.92 |
| Arrhythmia | 0.1343 | 0.0901 | 1.03 | (0.80, 1.31) | 0.84 |
| N follow-up days | 786 | 686 | | | |

**Note:** Results are derived from an inverse probability of treatment–weighted (IPTW) survival analysis evaluating the risk of new-onset secondary cardiovascular events among participants with baseline CVD in the intervention (INT) and control cohorts. Hazard ratios (HRs), 95% confidence intervals (CIs), and *p*-values were estimated using weighted Cox proportional hazards models adjusted for demographic and baseline covariates.
**Abbreviations:** CVD, cardiovascular disease; MACE-3, major adverse cardiac events (three-point composite endpoint); MACE-6, major adverse cardiac events (six-point composite endpoint); HR, hazard ratio; CI, confidence interval; INT, intervention; IPTW, inverse probability of treatment weighting.

**Appendix 8.** Multivariable Cox Regression Models Assessing Predictors of Cardiovascular and Hypertension Outcomes in the Intervention Cohort

| | **All CVD** | | **MACE-3** | | **MACE-6** | | **Hypertension** | |
|---|---|---|---|---|---|---|---|---|
| **Variables** | Hazard Ratio (95% CI) | p | Hazard Ratio (95% CI) | p | Hazard Ratio (95% CI) | p | Hazard Ratio (95% CI) | p |
| Ketone Ordinal Categories | 0.86 (0.77,0.96) | 0.01 | 1.05 (0.81, 1.37) | 0.71 | 0.98 (0.78, 1.23) | 0.85 | 1.09 (0.99, 1.20) | 0.06 |
| US Region: Northeast | 1.27 (0.87,1.85) | 0.21 | 1.07 (0.37, 3.1) | 0.9 | 1.06 (0.41, 2.74) | 0.91 | 1.45 (1.06, 1.99) | 0.02 |
| US Region: South | 1.29 (0.98,1.69) | 0.07 | 2.35 (1.17, 4.69) | 0.02 | 2.28 (1.25, 4.16) | 0.01 | 1.25 (1.00, 1.57) | 0.05 |
| US Region: West | 1.20 (0.83,1.72) | 0.33 | 1 (0.37, 2.67) | 1 | 1.84 (0.87, 3.90) | 0.11 | 1.37 (1.02, 1.83) | 0.04 |
| Age at index date | 1.07 (1.05,1.09) | <0.001 | 1.05 (1.01,1.09) | 0.01 | 1.03 (1.00, 1.06) | 0.05 | 0.99 (0.98, 1.00) | 0.29 |
| Female | Referent | 1 | Referent | 1 | Referent | 1 | Referent | 1 |
| Male | 1.47 (1.17,1.85) | <0.001 | 1.09 (0.63,1.88) | 0.76 | 1.24 (0.77, 1.98) | 0.38 | 1.02 (0.84, 1.24) | 0.81 |
| Missing Sex | 1.74 (0.55,5.50) | 0.35 | 2.8 (0.37,21) | 0.32 | 2.14 (0.29, 15.83) | 0.46 | 2.59 (1.14, 5.86) | 0.02 |
| White | Referent | 1 | Referent | 1 | Referent | 1 | Referent | 1 |
| Black or African American | 0.97 (0.66,1.43) | 0.88 | 1.66 (0.74,3.74) | 0.22 | 1.23 (0.61, 2.48) | 0.56 | 1.02 (0.74, 1.41) | 0.9 |
| Hispanic or Latino | 0.97 (0.63,1.51) | 0.91 | 1.95 (0.8,4.74) | 0.14 | 0.98 (0.43, 2.27) | 0.97 | 0.93 (0.65, 1.33) | 0.69 |
| Asian or Pacific Islander | 1.14 (0.63,2.08) | 0.67 | 1.53 (0.35,6.7) | 0.58 | 0.75 (0.18, 3.19) | 0.7 | 1.29 (0.84, 1.99) | 0.25 |
| Other Race | 2.18 (1.17,4.05) | 0.01 | 2.23 (0.51,9.66) | 0.28 | 1.28 (0.30, 5.39) | 0.74 | 0.64 (0.28, 1.44) | 0.28 |
| Missing Race | 1.1 (0.83,1.46) | 0.49 | 1.26 (0.62,2.54) | 0.52 | 0.90 (0.50, 1.62) | 0.72 | 0.95 (0.75, 1.19) | 0.64 |
| Baseline Obesity | 1.18 (0.94,1.49) | 0.15 | 0.92 (0.54,1.59) | 0.77 | 0.97 (0.61, 1.54) | 0.89 | 0.64 (0.53, 0.77) | 0 |
| Baseline Type 2 diabetes | 1.30 (1.00,1.67) | 0.05 | 2.52 (1.24, 5.14) | 0.01 | 2.57 (1.40, 4.72) | <0.001 | 0.98 (0.80, 1.20) | 0.84 |

**Note:** Hazard ratios (HRs), 95% confidence intervals (CIs), and *p*-values were estimated using Cox proportional hazards models. Each model included covariates for ketone ordinal categories, demographic factors (age, sex, and race/ethnicity), US region, baseline obesity, and baseline type 2 diabetes status. Analyses were restricted to participants in the intervention (INT) cohort.
**Abbreviations:** CVD, cardiovascular disease; MACE-3, major adverse cardiac events (three-point composite endpoint); MACE-6, major adverse cardiac events (six-point composite endpoint); HR, hazard ratio; CI, confidence interval.
